# Estimated morbimortality and costs attributable to child and adolescent obesity in Brazil from 2024 to 2060: a multistate life table study

**DOI:** 10.1101/2024.08.29.24312725

**Authors:** Eduardo Augusto Fernandes Nilson, Ana Carolina Rocha de Oliveira, Bruna Gusmão, Raphael Barreto da Conceição Barbosa

## Abstract

**Introduction:** Childhood obesity is a major global public health issue globally and in Brazil. The impacts of childhood obesity include higher risk of disease during childhood and of obesity and non-communicable diseases in adulthood and represent an important epidemiological and economic burden to countries.

**Methods:** This study is based on the multistate life table modeling of different scenarios of change3s in the child and adolescent obesity on the estimated costs attributable to obesity and the epidemiological burden of obesity-related diseases.

**Results:** According to this study, if the current trends in childhood and adolescent obesity in Brazil continue, the prevalence will significantly increase across different age subgroups and for both sexes by 2060 and obesity among adults will nearly double, resulting in R$3.84 billion costs attributable to childhood and adolescent obesity to the Brazilian Unified Health System during this period. Alternatively, if obesity prevalence among children and adolescents is reduced and remains constant these direct costs could be reduced by R$1.05 to R$1.27 billion by 2060 and up to 244,600 incident cases and 70,800 deaths from obesity-related diseases could be prevented.

**Conclusion:** This study highlights that the costs of childhood obesity are not limited to the impacts on adult health and represent a relevant economic burden to the Brazilian National Health System and to families because of additional costs during childhood. Therefore, the prevention and control of childhood obesity is a public health priority that demands immediate and robust policies.

## Introduction

Obesity among children and adolescents represents one of the greatest public health challenges today. Worldwide, childhood obesity has increased eightfold in the last four decades (1). According to the World Health Organization (WHO), in 2016, 340 million children and adolescents were overweight, and in 2020, 39 million children under the age of 5 were obese (2).

Considering consolidated analyses of cohort studies from 200 countries, different trends in height gain and variations in Body Mass Index (BMI) for age in children and adolescents were observed across different regions of the world. According to this study, in Brazil, cohorts of children and adolescents between 1985 and 2019 showed age-specific BMIs higher than WHO growth curves, consistent with the increasing trend in the prevalence of overweight and obesity among children and adolescents in the country (3).

In Brazil, data from national surveys show that the prevalence of obesity in children aged 5 to 9 years increased from 2.4% to 14.2% between 1974 and 2009, while the prevalence among adolescents increased from 0.6% to 5.0% in the same period (4). However, according to the Food and Nutrition Surveillance System (Sisvan) in 2022, the prevalence of obesity was 16.3% for children aged 5 to 9 years and 12.8% among adolescents monitored by Primary Health Care. The situation of childhood obesity is even more severe, as more recently, the National Survey of Child Nutrition (Enani), conducted in 2019, revealed that the prevalence of obesity in children under 5 years old was 10.1% (5). Growing evidence shows that, compared to eutrophic children, children who are overweight or obese are more likely to face issues such as poorer health even in childhood, including type 2 diabetes, asthma, hypertension, sleep apnea, musculoskeletal problems, and metabolic disorders (6), as well as lower self-esteem, a higher likelihood of being bullied, and lower school attendance and performance. Additionally, children and adolescents with obesity are more likely to have health problems in the future, as childhood obesity is a strong predictor of obesity in adulthood, increasing the risk of non-communicable chronic diseases such as type 2 diabetes, cardiovascular diseases, and certain types of cancer. The future impacts of childhood obesity also include a higher likelihood of negative socioeconomic and labor consequences, such as lower employability, reduced productivity, and lower wages (7).

Given this association between childhood and adolescent obesity and negative health outcomes in adulthood, many studies have begun to project the impacts of childhood obesity in terms of morbidity and mortality due to associated comorbidities, as well as direct costs for its treatment and indirect costs to society resulting from higher morbidity and mortality, early retirements, and productivity losses due to absenteeism and presenteeism (8)(9)(10)(11)(12).

Modeling studies of the impact of diseases and risk factors have been used to evaluate the impact of childhood obesity in other countries (13)(14), but they have not yet been developed for the Brazilian context. On the other hand, considering the adult population in Brazil, multi-state life table models have already been developed, from which it was possible to estimate the epidemiological burden of overweight and the associated direct and indirect costs. According to these studies, if the current trend of increasing overweight among adults continues, there will be approximately 5.26 million cases of diseases and 808.6 thousand deaths associated with overweight from 2021 to 2030, and the direct and indirect costs of these diseases will total around R$49.2 billion during this period (15)(16).

However, the costs of childhood obesity do not only occur in adulthood, and in a groundbreaking study in the country, it was estimated that the direct costs attributable to childhood obesity in the Unified Health System (SUS) reached approximately R$438.8 million in the last decade, including hospital, outpatient, and medication expenses (17). In Brazil, there are still no projections of the epidemiological and economic burden of obesity in children and adolescents, so this study aims to develop a multi-state life table model to estimate the growth trends of childhood obesity, the cases and deaths from diseases associated with obesity, and the direct and indirect costs of childhood obesity from 2024 to 2044.

## Methods

This study was based on the development and application of a multi-state life table model to estimate the future impacts of childhood and adolescent obesity in Brazil. The study was divided into the following stages: (1) projection of BMI evolution by age in children and adolescents and BMI in Brazilian adults from 2024 to 2060, assuming the continuation of current trends of average BMI increase for each age group; (2) projection of changes in BMI by age in children and adolescents and BMI in adults from 2024 to 2060 under different scenarios of childhood obesity reduction; (3) estimation of cases and deaths attributable to obesity, assuming the continuation of current trends of average BMI increase for each age group and under different scenarios of childhood obesity reduction from 2024 to 2060; (4) estimation of direct costs to the Unified Health System (SUS) attributable to childhood and adolescent obesity from 2024 to 2060; (5) estimation of direct and indirect costs of diseases and deaths attributable to obesity in Brazil that could be avoided under different scenarios of childhood obesity reduction from 2024 to 2060. The age groups of 5 to 9 years, 10 to 14 years, and 15 to 19 years were considered to represent children and adolescents based on the aggregation of age groups available in health information systems and national surveys.

### Estimation of Obesity Prevalence Trends

First, for the projection of BMI evolution by age in children and adolescents and BMI in Brazilian adults from 2024 to 2044, estimates obtained from the study by Rodriguez-Martinez et al. (2020) (18) were used to obtain average BMIs by sex and age and the trends in BMI growth by age in children and adolescents according to cohorts in Brazil from 1985 to 2019 using Prais Winsten regressions. The study by Estivaleti et al. (2022) (19) was used to obtain the trends in BMI growth in Brazilian adults according to projections from the Risk Factor Surveillance System for Chronic Disease Telephone Survey (Vigitel). The BMI information was then compiled and expanded in the multi-state life table model developed by Nilson et al. (2022) (15).

These trends in population obesity growth were then modeled for different scenarios of childhood obesity reduction in 2024 by 10% (most optimistic) and two scenarios with smaller reductions, 5% and 2%, and their impact on adult obesity until 2060. For the modeling, it was assumed that after the reduction in obesity prevalence in 2024, the same prevalence would be maintained among children and adolescents until 2060, and that the same rates of obesity prevalence growth in the population estimated by Estivaleti et al. (19) and extrapolated to 2060 would be maintained. Furthermore, the obesity growth trend was smoothed in the model considering the midpoint of each 5-year age range by sex through simple linear regression for each age group according to the usual scenario (current trends maintained in the future, or BAU – business as usual), extrapolated for each scenario of reducing childhood and adolescent obesity prevalence.

### Multi-State Life Table

The multi-state life table model assesses the association between overweight/obesity and eleven non-communicable chronic diseases (NCDs): ischemic heart disease, cerebrovascular diseases, hypertensive heart disease, diabetes, chronic kidney disease, liver cirrhosis, and cancers of the breast, colon, pancreas, kidney, and liver. The RR values for each NCD by age group were extracted to estimate the increased disease risk for each 5 kg/m² increase in BMI. Data on incidence, prevalence, and mortality from NCDs were extracted from the Global Burden of Disease (GBD) study (20), stratified by sex and age group (considering 5-year age ranges), identified by the International Classification of Diseases 10 (ICD-10) codes, and data on case fatality and remission were estimated using the Dismod II tool (21).

The cohort simulation model by multi-state life table (MSLT) of cases and deaths from 11 NCDs was adapted from another validated study for estimating NCDs attributable to overweight in Brazil (Nilson et al., 2022). The model considers a hypothetical cohort of individuals who, in each cycle (year), are classified as healthy, ill, or deceased based on the incidence, mortality, remission, and case fatality rates of the diseases considered in the study. For this, within each sex and age stratum and for each disease evaluated, the RR values and the prevalence of overweight and non-communicable chronic diseases were considered, through systematic reviews and meta-analyses. The BMI distribution in the population was modeled considering a log-linear function of the average BMI and its standard deviation and the national population for each sex and age stratum.

Finally, within each sex and age stratum, for the scenario of maintaining BMI increase trends and for each reduction scenario, we calculated the potential impact fraction (PIF) for outcomes (o) in each age group (a) and sex (s). The PIF results represent the percentage difference in incidence and mortality rates of NCDs attributable to scenarios of population BMI increase. The model assumes that the impact of obesity reduction on NCD incidence and mortality rates would not occur immediately, so latency periods were incorporated into the model between BMI changes and outcomes, considering an average of 5 years for cardiovascular diseases, type 2 diabetes, and chronic kidney disease, and an average of 10 years for cancers. The uncertainty of cases and deaths attributable to different scenarios was quantified through Monte Carlo simulations (n = 5,000), where for each simulation, a draw was made from the distributions (uncertainties) of BMI and RR, which were reported for the 50th, 2.5th, and 97.5th percentiles of estimates across all simulations as the central estimate and 95% uncertainty intervals, respectively.

### Estimates of Direct Costs attributable to Obesity

For estimating the direct costs to the Unified Health System (SUS) attributable to childhood and adolescent obesity until 2044, the same methodology by Nilson et al. (2023) (22) was used, which estimates excess costs for hospitalizations of children and adolescents with obesity compared to those with an adequate BMI, using data from the SUS Hospital Information System (SIH/SUS), using the base year of 2022. Next, to estimate non-hospital, outpatient, and medication expenses, it was assumed that these would be proportional to additional hospitalization costs according to the meta-analysis by Ling et al., 2023 (23).

For estimating direct costs in adulthood, the cost data for NCDs in the SUS were extracted using the Tabnet tool for hospitalizations (Hospital Information System – SIH/SUS) and outpatient care (Outpatient Information System – SIA/SUS), publicly available from the SUS Department of Informatics (DATASUS). Costs were extracted by sex and age group (considering 5-year age ranges) and for the 11 diseases included in the multi-state life table model. To calculate the per capita cost of NCDs in the SUS, the costs from SIH and SIA were used, considering the average cost per patient for the SUS. This average cost per patient (numerator) was multiplied by the number of incident cases attributable to obesity until 2044.

Further details of the parameters for the estimations and the modeling methodology are described in the Supplementary Materials.

## Results

*Future Trends in Childhood and Adolescent Obesity prevalences and direct costs* Following the trends observed in Brazilian cohorts from 1985 to 2019, estimates show that obesity prevalence will increase across all age groups for both sexes until 2060. From 2023 to 2060, among boys aged 5 to 9 years, obesity prevalence will rise from 22.2% to 34.7%, while it will increase from 14.3% to 24.9% among girls of the same age. In the same period, obesity prevalence will increase from 12.6% to 21.9% among boys aged 10 to 14 years and from 8.3% to 15.5% among girls in this age group. Finally, in the next two decades, for the 15 to 19 age group, obesity prevalence among boys will rise from 8.6% to 16.0% and from 7.9% to 14.9% among girls.

Following the same methodology adopted in the previous study on direct costs of childhood and adolescent obesity, we used the annual per capita costs in 2022 as a reference to estimate hospital, non-hospital, outpatient, and medication costs attributable to obesity in individuals aged 5 to 19 years in Brazil from 2013 to 2022.

If current trends in prevalence increases continue, the annual direct attributable costs would rise from R$83.2 million in 2023 to R$115.5 million in 2060, totaling R$3.84 billion over the period. Most of these costs (48.1%) would be in the 15 to 19 age group, while 18.8% would occur in the 10 to 14 age group, and 33.2% of these costs would be among individuals aged 5 to 9 years. In terms of cost distribution by type, 94.4% would be spent on hospital expenses, 2.7% on non-hospital expenses, 2.2% on medications, and 0.7% on outpatient care and procedures (Table 1).

**Table 1.**
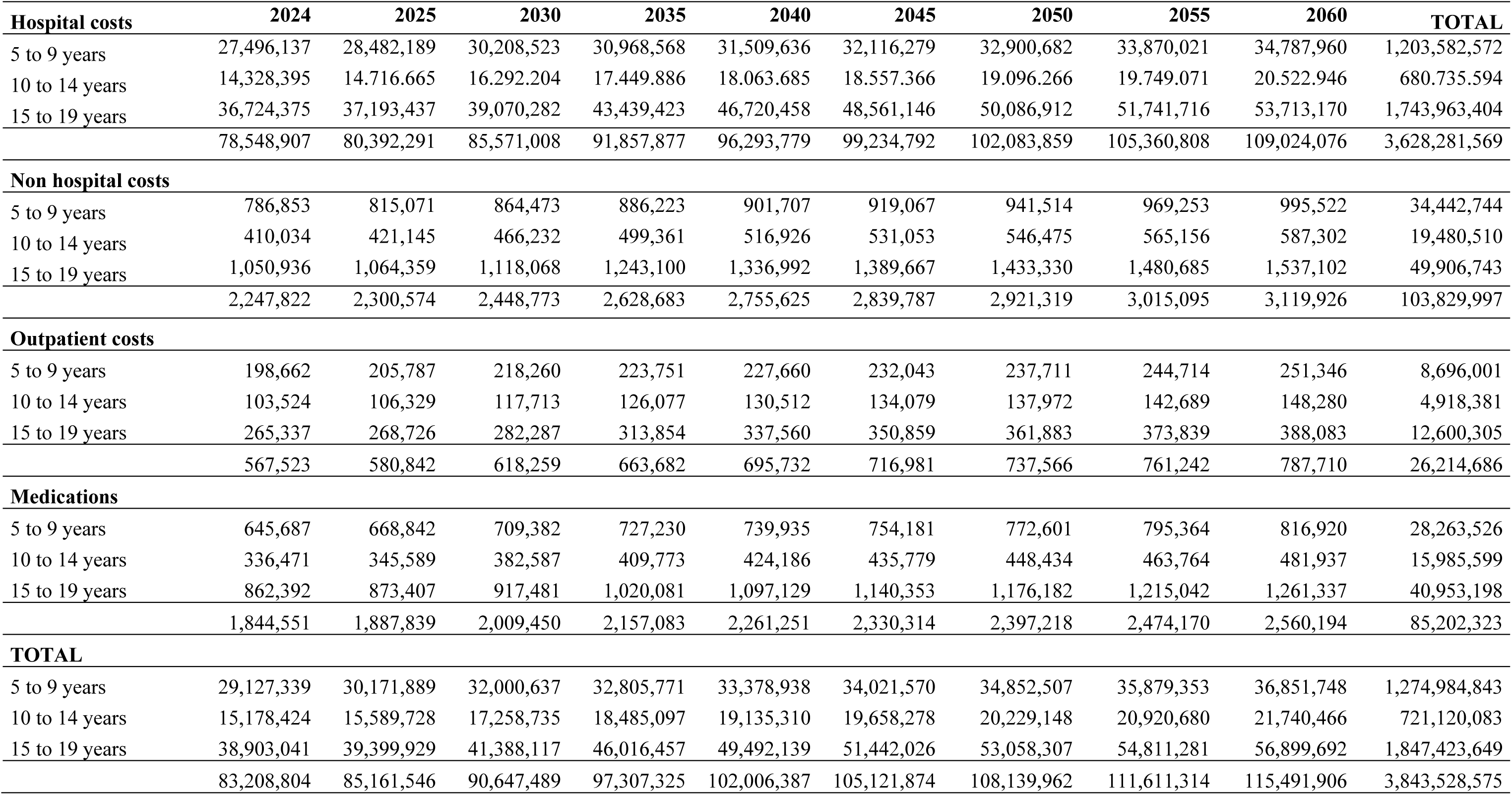
Estimates of the future direct costs attributable to child and adolescent obesity from2024 to 2060 in Brazil (in Brazilian Reals – R$).

| **Hospital costs** | **2024** | **2025** | **2030** | **2035** | **2040** | **2045** | **2050** | **2055** | **2060** | **TOTAL** |
| --- | --- | --- | --- | --- | --- | --- | --- | --- | --- | --- |
| 5 to 9 years | 27,496,137 | 28,482,189 | 30,208,523 | 30,968,568 | 31,509,636 | 32,116,279 | 32,900,682 | 33,870,021 | 34,787,960 | 1,203,582,572 |
| 10 to 14 years | 14,328,395 | 14,716,665 | 16,292,204 | 17,449,886 | 18,063,685 | 18,557,366 | 19,096,266 | 19,749,071 | 20,522,946 | 680,735,594 |
| 15 to 19 years | 36,724,375 | 37,193,437 | 39,070,282 | 43,439,423 | 46,720,458 | 48,561,146 | 50,086,912 | 51,741,716 | 53,713,170 | 1,743,963,404 |
|  | 78,548,907 | 80,392,291 | 85,571,008 | 91,857,877 | 96,293,779 | 99,234,792 | 102,083,859 | 105,360,808 | 109,024,076 | 3,628,281,569 |
| **Non hospital costs** |  |  |  |  |  |  |  |  |  |  |
| 5 to 9 years | 786,853 | 815,071 | 864,473 | 886,223 | 901,707 | 919,067 | 941,514 | 969,253 | 995,522 | 34,442,744 |
| 10 to 14 years | 410,034 | 421,145 | 466,232 | 499,361 | 516,926 | 531,053 | 546,475 | 565,156 | 587,302 | 19,480,510 |
| 15 to 19 years | 1,050,936 | 1,064,359 | 1,118,068 | 1,243,100 | 1,336,992 | 1,389,667 | 1,433,330 | 1,480,685 | 1,537,102 | 49,906,743 |
|  | 2,247,822 | 2,300,574 | 2,448,773 | 2,628,683 | 2,755,625 | 2,839,787 | 2,921,319 | 3,015,095 | 3,119,926 | 103,829,997 |
| **Outpatient costs** |  |  |  |  |  |  |  |  |  |  |
| 5 to 9 years | 198,662 | 205,787 | 218,260 | 223,751 | 227,660 | 232,043 | 237,711 | 244,714 | 251,346 | 8,696,001 |
| 10 to 14 years | 103,524 | 106,329 | 117,713 | 126,077 | 130,512 | 134,079 | 137,972 | 142,689 | 148,280 | 4,918,381 |
| 15 to 19 years | 265,337 | 268,726 | 282,287 | 313,854 | 337,560 | 350,859 | 361,883 | 373,839 | 388,083 | 12,600,305 |
|  | 567,523 | 580,842 | 618,259 | 663,682 | 695,732 | 716,981 | 737,566 | 761,242 | 787,710 | 26,214,686 |
| **Medications** |  |  |  |  |  |  |  |  |  |  |
| 5 to 9 years | 645,687 | 668,842 | 709,382 | 727,230 | 739,935 | 754,181 | 772,601 | 795,364 | 816,920 | 28,263,526 |
| 10 to 14 years | 336,471 | 345,589 | 382,587 | 409,773 | 424,186 | 435,779 | 448,434 | 463,764 | 481,937 | 15,985,599 |
| 15 to 19 years | 862,392 | 873,407 | 917,481 | 1,020,081 | 1,097,129 | 1,140,353 | 1,176,182 | 1,215,042 | 1,261,337 | 40,953,198 |
|  | 1,844,551 | 1,887,839 | 2,009,450 | 2,157,083 | 2,261,251 | 2,330,314 | 2,397,218 | 2,474,170 | 2,560,194 | 85,202,323 |
| **TOTAL** |  |  |  |  |  |  |  |  |  |  |
| 5 to 9 years | 29,127,339 | 30,171,889 | 32,000,637 | 32,805,771 | 33,378,938 | 34,021,570 | 34,852,507 | 35,879,353 | 36,851,748 | 1,274,984,843 |
| 10 to 14 years | 15,178,424 | 15,589,728 | 17,258,735 | 18,485,097 | 19,135,310 | 19,658,278 | 20,229,148 | 20,920,680 | 21,740,466 | 721,120,083 |
| 15 to 19 years | 38,903,041 | 39,399,929 | 41,388,117 | 46,016,457 | 49,492,139 | 51,442,026 | 53,058,307 | 54,811,281 | 56,899,692 | 1,847,423,649 |
|  | 83,208,804 | 85,161,546 | 90,647,489 | 97,307,325 | 102,006,387 | 105,121,874 | 108,139,962 | 111,611,314 | 115,491,906 | 3,843,528,575 |

### Scenarios for Reducing Childhood and Adolescent Obesity

If current trends in obesity prevalence growth continue across all age groups in Brazil, the number of people aged 5 to 79 with obesity will rise from 26.7 million in 2023 to 38.6 million in 2060, representing a 44% increase. In terms of sex and age groups, this corresponds to 4.7 million boys and 2.8 million girls with obesity in 2023, and 5.9 million boys and 3.8 million girls with obesity in 2060 (an increase of 22% to 36%). Among adults, it is estimated that there would be 10.1 million men and 9.2 million women with obesity in 2023, and if the current trends in prevalence growth continue, these numbers would rise to 15.4 million men and 13.8 million women in 2060 (increases of 50% to 52%).

Considering the scenarios for reducing childhood and adolescent obesity, a 10% reduction in obesity prevalence among individuals aged 5 to 19 would slow the growth in adult obesity prevalence among those up to 59 years old. While in the baseline scenario (BAU – business-as-usual), which reflects the continuation of current trends, there would be an average 99.5% increase in obesity prevalence among adults (ranging from 86.5% to 148%, depending on sex and age group), in the scenario with a 10% reduction in obesity prevalence among children and adolescents, the average increase in adult obesity prevalence would be 17.8% (ranging from 1.2% to 72.6%). In the intermediate scenarios, the average increase in obesity prevalence among adults up to 59 years old would decrease to 23.4% for a 5% reduction in childhood and adolescent obesity and to 26.8% for a 2% reduction, with ranges from 6.8% to 75.0% and from 10.2% to 76.4%, respectively (Tables 2 and 3).

**Table 2.** Estimated prevalences according to the maintenance of current trends and in different scenarios of reduction of the prevalence of ity among children and adolescents in Brazil from 2024 to 2060.

|  | Current trends |  | 10% reduction |  | 5% reduction |  | 2% reduction |  |
| --- | --- | --- | --- | --- | --- | --- | --- | --- |
|  | 2024 | 2060 | 2024 | 2060 | 2024 | 2060 | 2024 | 2060 |
| **Men** |  |  |  |  |  |  |  |  |
| **20 to 24 years** | 15.34% | 30.85% | 15.34% | 15.54% | 15.34% | 16.41% | 15.34% | 16.93% |
| **25 to 29 years** | 29.49% | 59.25% | 29.49% | 31.51% | 29.49% | 33.26% | 29.49% | 34.31% |
| **30 to 34 years** | 36.82% | 69.18% | 36.82% | 42.09% | 36.82% | 44.43% | 36.82% | 45.83% |
| **35 to 39 years** | 37.37% | 69.88% | 37.37% | 43.00% | 37.37% | 45.38% | 37.37% | 46.82% |
| **40 to 44 years** | 37.70% | 70.83% | 37.70% | 42.87% | 37.70% | 45.25% | 37.70% | 46.68% |
| **45 to 49 years** | 36.12% | 69.84% | 36.12% | 40.17% | 36.12% | 42.40% | 36.12% | 43.74% |
| **50 to 54 years** | 36.27% | 71.08% | 36.27% | 40.21% | 36.27% | 42.44% | 36.27% | 43.78% |
| **54 to 59 years** | 34.08% | 68.78% | 34.08% | 56.29% | 34.08% | 57.12% | 34.08% | 57.61% |
| **Women** |  |  |  |  |  |  |  |  |
| **20 to 24 years** | 24.37% | 40.09% | 24.37% | 24.67% | 24.37% | 26.04% | 24.37% | 26.86% |
| **25 to 29 years** | 31.99% | 61.08% | 31.99% | 34.11% | 31.99% | 36.01% | 31.99% | 37.14% |
| **30 to 34 years** | 34.38% | 65.00% | 34.38% | 39.30% | 34.38% | 41.49% | 34.38% | 42.80% |
| **35 to 39 years** | 35.58% | 66.93% | 35.58% | 40.94% | 35.58% | 43.21% | 35.58% | 44.57% |
| **40 to 44 years** | 36.21% | 67.48% | 36.21% | 41.16% | 36.21% | 43.44% | 36.21% | 44.81% |
| **45 to 49 years** | 36.82% | 69.90% | 36.82% | 40.96% | 36.82% | 43.23% | 36.82% | 44.60% |
| **50 to 54 years** | 32.22% | 67.46% | 32.22% | 35.71% | 32.22% | 37.70% | 32.22% | 38.89% |
| **54 to 59 years** | 30.83% | 65.41% | 30.83% | 53.23% | 30.83% | 53.96% | 30.83% | 54.40% |

**Table 3.** Estimates of the predicted number of individuals with obesity in Brazil from 2024 to 2060 by sex and age group.

|  | 2024 | 2025 | 2030 | 2035 | 2040 | 2045 | 2050 | 2055 | 2060 |
| --- | --- | --- | --- | --- | --- | --- | --- | --- | --- |
| **Men** |  |  |  |  |  |  |  |  |  |
| **5 to 9 years** | 1,677,255 | 1,717,557 | 1,785,681 | 1,812,848 | 1,835,969 | 1,866,307 | 1,907,812 | 1,955,494 | 1,992,756 |
| **10 to 14 years** | 1,554,238 | 1,606,763 | 1,682,035 | 1,713,656 | 1,737,900 | 1,767,555 | 1,807,421 | 1,855,549 | 1,897,024 |
| **15 to 19 years** | 1,444,340 | 1,493,108 | 1,578,791 | 1,614,732 | 1,639,954 | 1,668,819 | 1,706,930 | 1,754,372 | 1,798,582 |
| **20 to 24 years** | 1,274,297 | 1,316,212 | 1,410,170 | 1,449,871 | 1,476,604 | 1,505,084 | 1,541,536 | 1,587,368 | 1,632,998 |
| **25 to 29 years** | 1,088,077 | 1,136,473 | 1,224,829 | 1,271,932 | 1,300,484 | 1,328,935 | 1,363,950 | 1,407,695 | 1,453,807 |
| **30 to 34 years** | 939,068 | 982,464 | 1,067,391 | 1,120,514 | 1,150,693 | 1,179,091 | 1,212,649 | 1,254,008 | 1,299,662 |
| **35 to 39 years** | 801,436 | 829,233 | 917,650 | 972,956 | 1,004,926 | 1,033,302 | 1,065,430 | 1,104,290 | 1,148,689 |
| **40 to 44 years** | 694,657 | 715,541 | 796,111 | 855,486 | 888,967 | 917,005 | 947,503 | 983,810 | 1,026,209 |
| **45 to 49 years** | 671,262 | 686,832 | 758,640 | 826,514 | 862,401 | 890,514 | 919,834 | 954,440 | 995,485 |
| **50 to 54 years** | 664,353 | 673,581 | 740,699 | 809,518 | 852,158 | 880,787 | 909,249 | 942,433 | 982,084 |
| **55 to 59 years** | 662,109 | 670,896 | 727,927 | 799,725 | 848,921 | 878,535 | 906,458 | 938,376 | 976,580 |
| **60 to 64 years** | 693,458 | 706,958 | 756,833 | 850,002 | 913,459 | 953,983 | 989,968 | 1,028,776 | 1,073,558 |
| **65 to 69 years** | 763,991 | 782,429 | 841,448 | 956,642 | 1,051,392 | 1,113,445 | 1,166,320 | 1,220,212 | 1,279,658 |
| **70 to 74 years** | 844,149 | 866,579 | 943,875 | 1,081,590 | 1,223,831 | 1,318,163 | 1,396,909 | 1,473,621 | 1,554,973 |
| **75 to 79 years** | 1,015,269 | 1,040,802 | 1,138,631 | 1,306,845 | 1,492,222 | 1,632,757 | 1,737,071 | 1,834,041 | 1,933,642 |

(Table 3 – continuation)
|  | 2024 | 2025 | 2030 | 2035 | 2040 | 2045 | 2050 | 2055 | 2060 |
| --- | --- | --- | --- | --- | --- | --- | --- | --- | --- |
| **Women** |  |  |  |  |  |  |  |  |  |
| **5 to 9 years** | 1,029,984 | 1,062,418 | 1,120,224 | 1,154,680 | 1,187,167 | 1,224,751 | 1,270,119 | 1,319,932 | 1,362,934 |
| **10 to 14 years** | 928,074 | 966,314 | 1,025,906 | 1,061,037 | 1,092,302 | 1,127,446 | 1,169,592 | 1,217,484 | 1,261,319 |
| **15 to 19 years** | 813,293 | 846,719 | 909,120 | 944,677 | 974,623 | 1,007,213 | 1,045,885 | 1,090,843 | 1,134,303 |
| **20 to 24 years** | 723,421 | 752,072 | 819,265 | 855,574 | 884,492 | 914,749 | 950,282 | 992,209 | 1,034,797 |
| **25 to 29 years** | 639,156 | 670,947 | 733,857 | 772,911 | 800,690 | 828,660 | 861,110 | 899,601 | 940,374 |
| **30 to 34 years** | 589,685 | 618,543 | 679,961 | 721,878 | 749,021 | 775,221 | 805,139 | 840,645 | 879,677 |
| **35 to 39 years** | 542,192 | 561,917 | 627,732 | 671,414 | 699,278 | 724,754 | 753,087 | 786,502 | 824,400 |
| **40 to 44 years** | 529,436 | 545,798 | 610,056 | 658,955 | 688,341 | 713,555 | 740,835 | 772,863 | 809,988 |
| **45 to 49 years** | 519,423 | 531,613 | 589,387 | 645,313 | 676,631 | 701,860 | 728,159 | 758,828 | 794,903 |
| **50 to 54 years** | 551,309 | 558,908 | 615,121 | 674,316 | 712,436 | 738,527 | 764,552 | 794,673 | 830,431 |
| **55 to 59 years** | 586,472 | 594,139 | 643,692 | 708,052 | 753,106 | 780,498 | 806,429 | 835,996 | 871,247 |
| **60 to 64 years** | 709,509 | 723,594 | 769,367 | 862,698 | 923,219 | 959,316 | 989,351 | 1,021,255 | 1,058,453 |
| **65 to 69 years** | 929,183 | 949,884 | 1,008,210 | 1,132,458 | 1,230,156 | 1,283,636 | 1,322,169 | 1,359,187 | 1,400,589 |
| **70 to 74 years** | 1,256,785 | 1,277,344 | 1,358,874 | 1,516,134 | 1,675,318 | 1,755,987 | 1,807,987 | 1,852,651 | 1,900,099 |
| **75 to 79 years** | 1,602,398 | 1,621,787 | 1,723,759 | 1,914,564 | 2,120,283 | 2,244,384 | 2,309,785 | 2,360,394 | 2,411,380 |

These impacts illustrate the combined effect of the initial reduction in childhood and adolescent obesity prevalence, coupled with its maintenance over time, ensuring that these impacts gradually extend to adults in the future, following the cohort in the life table. For example, reductions among individuals aged 15 to 19 will first impact younger adults (aged 20 to 24) in the first five years of the simulation. Over time, more adult age groups will be affected, eventually reaching those aged 55 to 59 by 2060.

### Impact of Reduction Scenarios on the Direct Costs Attributable to Childhood and Adolescent Obesity

In terms of the potential impact of reducing and subsequently maintaining the prevalence of obesity among children and adolescents, significant cost savings could be achieved in all scenarios when considering the costs attributable to childhood and adolescent obesity within the SUS. In the most optimistic scenario, with a 10% reduction in prevalence, approximately R$1.274 billion could be saved by 2060, while a 5% reduction would lead to savings of R$1.136 billion, and a 2% reduction would result in savings of R$1.053 billion over the same period (Table 4).

**Table 4.** Estimated savings in the direct health costs attributable to child and adolescent obesity from 2024 to 2060 in different scenarios of prevalence reduction in Brazil (in Brazilian Reals – R$).

| **TOTAL** | **10%** | **5%** | **2%** |
| --- | --- | --- | --- |
| 5 to 9 years | 382,196,079 | 334,214,889 | 305,426,175 |
| 10 to 14 years | 240,655,798 | 214,806,583 | 199,297,055 |
| 15 to 19 years | 651,362,302 | 587,075,730 | 548,503,786 |
|  | **1,274,214,180** | **1,136,097,203** | **1,053,227,016** |

**Table 5.** Prevented or averted incident cases of obesity-related diseases from 2024 to 2060 in different scenarios of obesity reduction among ren and adolescents in Brazil (Mean and Uncertainty Intervals 95%).

| **Scenario** | **Incident cases** | **Deaths** |
| --- | --- | --- |
| **10%** |  |  |
| Men | 74,227 (44,818-101,972) | 26,298 (13,391-37,486) |
| Women | 170,034 (80,730-244,107) | 44,542 (17,366-64,952) |
| Total | 244,261 (125,547-346,079) | 70,840 (30,757-102,438) |
| **5%** |  |  |
| Men | 65,847 (39,744-90,476) | 23,371 (11,899-33,319) |
| Women | 151,732 (72,052-217,867) | 39,817 (15,523-58,072) |
| Total | 217,579 (111,796-308,343) | 63,189 (27,422-91,391) |
| **2%** |  |  |
| Men | 60,937 (36,772-83,738) | 21,651 (11,022-30,869) |
| Women | 140,996 (66,963-202,469) | 37,038 (14,439-54,024) |
| Total | 201,932 (103,734-286,207) | 58,689 (25,461-84,893) |

**Table 6.** Incident cases of obesity related diseases prevented from 2024 to 2060 in different scenarios of obesity reduction among children and adolescents by disease group in Brazil (Mean and Uncertainty Intervals 95%).

|  | Prevented cases |  |  |
| --- | --- | --- | --- |
|  | 2% | 5% | 10% |
| Ischaemic heart disease | 7,743 (3,623-11,298) | 8,320 (3,894-12,139) | 9,294 (4,351-13,560) |
| Cerebrovascular diseases | 23,959 (11,792-34,629) | 25,779 (12,687-37,259) | 28,873 (14,210-41,730) |
| Hypertensive heart disease | 17,24 (832-2,391) | 1,857 (896-2,576) | 2,085 (1,006-2,893) |
| Diabetes | 116,268 (64,109-157,003) | 125,451 (69,174-169,397) | 141,167 (77,843-190,611) |
| Cancers | 136 (73-207) | 147 (79-224) | 165 (89-251) |
| Cirrhosis | 11,715 (6,464-16,608) | 12,627 (6,968-17,901) | 14,178 (7,826-20,099) |
| Chronic kidney disease | 40,388 (16,842-64,070) | 43,399 (18,097-68,846) | 48,499 (20,224-76,935) |
| Total | 201,932 (103,734-286,207) | 217,579 (111,796-308,343) | 244,261 (125,547-346,079) |

**Table 7.** Deaths of obesity related diseases prevented from 2024 to 2060 in different scenarios of obesity reduction among children and adolescents by disease group in Brazil (Mean and Uncertainty Intervals 95%).

| **Deaths** |  |  |  |
| --- | --- | --- | --- |
|  | **2%** | **5%** | **10%** |
| Ischaemic heart disease | 7,767 (3,239-11,404) | 8,345 (3,239-11,404) | 9,322 (3,890-13,687) |
| Cerebrovascular diseases | 15,072 (5,991-23,457) | 16,218 (5,991-23,457) | 18,164 (7,223-28,266) |
| Hypertensive heart disease | 1,769 (766-2,447) | 1,906 (766-2,447) | 2,141 (927-2,960) |
| Diabetes | 15,281 (7,136-19,741) | 16,480 (7,136-19,741) | 18,531 (8,658-23,941) |
| Cancers | 134 (63-206) | 144 (63-206) | 162 (76-250) |
| Cirrhosis | 10,900 (5,219-15,382) | 11,751 (5,219-15,382) | 13,197 (6,323-18,623) |
| Chronic kidney disease | 7,767 (3,047-12,256) | 8,345 (3,047-12,256) | 9,322 (3,660-14,710) |
| Total | 58,689 (25,461-84,893) | 63,189 (25,461-84,893) | 70,840 (30,757-102,438) |

## DISCUSSION

According to this study, if the current trends in childhood and adolescent obesity in Brazil continue, the prevalence will significantly increase across different age subgroups and for both sexes by 2060, potentially nearly doubling among children aged 10 to 14 and adolescents. As a result, the costs attributable to childhood and adolescent obesity to the SUS (Unified Health System) could reach R$3.84 billion during this period. On the other hand, if obesity prevalence among children and adolescents is reduced and remains constant, these direct costs could be reduced by R$1.05 to R$1.27 billion by 2060.

Additionally, considering the impact of childhood and adolescent obesity on adulthood, if the trends continue until 2060, obesity among adults will nearly double. However, in scenarios where obesity prevalence in childhood and adolescence is reduced and then maintained, there would be a significant slowdown in the rate of obesity increase among adults. As a consequence of this reduction, across different scenarios studied (2% to 10% reduction in childhood and adolescent obesity prevalence), up to 244,600 incident cases and 70,800 deaths from obesity-related diseases could be prevented. Among the outcomes, cardiovascular diseases, diabetes, and chronic kidney disease had the greatest impact on the prevented cases and deaths due to reduced obesity prevalence in childhood and adolescence.

These estimates demonstrate that reducing childhood and adolescent obesity is crucial to lowering adult obesity and its consequences on the future morbidity and mortality of Brazilians. Moreover, the impact on direct health costs is likely to grow even more in the future if current trends continue, leading to a significant increase in direct and indirect costs attributable to obesity in the adult population in the future.

Regarding the costs to the SUS attributable to obesity, it should be noted that they already reached R$1.42 million in 2018, and future projections indicate that the direct and indirect costs of obesity could reach 6.09% of Brazil’s Gross Domestic Product, approximately US$243.23 billion.

If current trends in the growth of obesity and overweight among the Brazilian adult population continue, their prevalence will reach 29.6% and 68.1%, respectively, by 2030. During this period, 5.26 million new cases and 808,000 deaths from cardiovascular diseases, cancers, diabetes, chronic kidney diseases, among others, will be attributable to excess weight by the end of this decade. Additionally, another study using the same simulation model applied in this research estimated that, between 2020 and 2030, around R$4.2 billion would be spent on direct expenses of NCDs if the current trend in the prevalence of excess weight among adults in Brazil continues, with indirect costs associated with premature mortality reaching approximately R$45.5 billion in the same period.

These changes in obesity prevalence across all age groups, including children and adolescents, are strongly associated with the nutritional transition that Brazil is undergoing, with the gradual replacement of the traditional diet—composed of rice, beans, cereals, fruits, vegetables, and other food types—by ultra-processed products in all age groups, including children and adolescents. According to the National Study on Child Nutrition (Enani 2019), about 80% of children under 2 years old consumed ultra-processed products, despite the Ministry of Health’s recommendation against this practice. It is even more concerning that, according to the 2017-2018 Household Budget Survey by the Brazilian Institute of Geography and Statistics (POF/IBGE), the highest proportion of ultra-processed foods in total dietary energy is among adolescents (reaching 26.7% of dietary calories) compared to adults and the elderly.

A growing body of scientific evidence reinforces that the consumption of ultra-processed products is directly related to a higher risk of overweight and obesity, as well as other noncommunicable diseases such as cardiovascular diseases, diabetes, and certain types of cancer. In addition to studies based on cohorts and surveys, and meta-analyses derived from them, a recent randomized clinical trial demonstrated that a diet based on ultra-processed foods compared to a diet based on whole and minimally processed foods is associated with higher calorie intake and weight gain.

Childhood obesity is recognized as a risk factor for obesity in adulthood, with 55% of children with obesity likely to become adolescents with obesity, and about 80% of adolescents with obesity expected to have obesity by the age of 30. Furthermore, childhood obesity is associated with a moderate increase in the risk of morbidity in adulthood, including diabetes, cardiovascular diseases (such as ischemic heart disease and hypertension), and some types of cancer.

Moreover, childhood obesity is associated with higher healthcare costs, including hospitalizations, outpatient costs, medication expenses, and other costs to families, as well as longer hospital stays for various diseases exacerbated by obesity, which also entails significant direct and indirect costs among adults. In Brazil, the costs attributable to childhood and adolescent obesity for the treatment of diseases within the SUS up to the age of 19 exceed the total costs of hospitalizations for diseases classified under Chapter V of the ICD-10, which corresponds to endocrine, nutritional, and metabolic diseases.

The current study is based on methodologies already validated in other studies on the projection of the impact of childhood and adolescent obesity on adulthood, and the current adaptation of a multi-state life table model for the Brazilian population from age 5 onwards could serve as a useful tool for new studies on the burden of obesity-related diseases and for modeling the potential impact of policies and interventions on health and nutrition in childhood, adolescence, and adulthood, as well as evaluating their costs.

The results highlight the significant epidemiological and economic impact of childhood and adolescent obesity in Brazil, emphasizing the need and urgency for more cost-effective policies to treat overweight and obesity within the SUS from childhood, along with the adoption of regulatory and fiscal policies that promote healthier food environments throughout the life cycle, especially in childhood and adolescence. These nutrition protection measures begin with the promotion and protection of breastfeeding and complementary feeding and continue through the regulation of food advertising, the regulation of ultra-processed food sales in schools, the improvement of nutritional labeling, and the taxation of ultra-processed products, along with subsidies for whole and minimally processed foods, as well as the strengthening of food and nutrition education actions for the population based on the Brazilian Dietary Guidelines and the Dietary Guidelines for Children Under 2 Years Old.

As strengths of this study, the direct cost estimation of obesity was based on modeling from a recent and robust meta-analysis on the costs of childhood and adolescent obesity worldwide and hospital expenses of the Brazilian public health system. For the prevalence of childhood and adolescent obesity, published data from a set of Brazilian cohorts were used, covering nutritional data for individuals aged 5 to 19 years up to 2019, including to model future trends in weight gain among children and adolescents. Finally, the modeling of the impact of changes in childhood obesity on adult health outcomes was adapted from a validated and published multi-state life table model. Additionally, the methods adapted and developed in this study could serve as a basis for new analyses and projections, including the potential impact of policies to reduce obesity among children and adolescents.

Among its limitations, we have the adoption of hospital cost estimates from primary studies in other countries, with different demographic, socioeconomic characteristics, and health systems compared to Brazil, possible limitations in the actual expenditures within the SUS, the assumptions adopted in incorporating the cohort’s nutritional data into the life table model, as well as the limitations already described regarding the multi-state life table for Brazil.

However, the assumptions adopted in the various parts of this study aimed to generate more conservative estimates for the outcomes, while also incorporating uncertainty analysis through Monte Carlo analysis for the projections of cases and deaths attributable in adulthood. Future analyses should further consider the economic evaluation of the impact of childhood and adolescent obesity and analyze new scenarios of changes in prevalence, as well as expand the model to years beyond 2060 when new population projections are generated by the IBGE.

In conclusion, the results of this study add to those of the previous study of this project, reinforcing that childhood obesity is a major public health problem and a significant epidemiological and economic issue for Brazilian society, considering its current and future impacts if nothing is done. Therefore, immediate and effective measures are essential to address it, including treatment and prevention policies.

## Data Availability

All relevant data on the model are publicly available. Hospitalization data is available from http://tabnet.datasus.gov.br/cgi/deftohtm.exe?sih/cnv/niuf.def and IHME

## Declarations

Ethics approval and consent to participate Not applicable.

### Authors’ contributions

EAFN conceptualised the idea and was responsible for the study design and data analysis and drafted the original manuscript. All authors revised and approved the final manuscript.

