## Supplementary material for "Estimated morbimortality and costs attributable to child and adolescent obesity in Brazil from 2024 to 2060: a multistate life table study": https://github.com/eduardonilson/Obesity-MSLT-BR/blob/main/Supplementary%20materials%20MSLT%20children%20and%20adolescents.pdf (from another version of this preprint: version 1, 2024-08-30)

### Appendix – Supplementary Materials

*Estimation of the number of hospitalizations according to BMI by age-group*

We used the prevalence of children and adolescents with obesity and those with adequate BMI and conservatively assumed that both groups have similar risk of hospitalization from all-causes.

We used the following formula for the estimation:

$$Int_t = Int_t * P_{Ob} + Int_t * P_{NOb}$$

Onde:

$Int_t$ = total number of hospitalizations

$P_{Ob}$ = prevalence of children and adolescents with obesity

$P_{NOb}$ = prevalence of children and adolescents without obesity

*Estimation of the average per capita costs of hospitalizations by nutritional status*

Firstly, we estimated the average per capita cost of hospitalizations for each group (children and adolescents with and without obesity), considering the estimated percentage of additional hospitalization costs.

Finally, the average costs of hospitalizations were multiplied by the total number of hospitalizations for the total costs of hospitalizations and the difference between the average per capita costs of hospitalizations of children and adolescents with and without obesity was used to calculate the additional costs attributable to childhood obesity.

*Proportion of non-hospital, outpatient and medication costs*

The results of the meta-anlaysis by Ling et al (1), was used to estimate the proportion of non-hospital, outpatient and medication costs in relation to the hospitalization costs and used these proportions to multiply the total and additional costs (Table S6), assuming that the same proportionality would be applied to the Brazailian population.

**Table S6. Proportion of non-hospital, outpatient and medication costs in relation to the hospitalization costs from Ling et al, 2023.**

| Non-hospital costs | Outpatient costs | Medication costs |
|---|---|---|
| 2.86% | 0.80% | 2.35% |

**Multistate life table model**

This proposed model estimates the effect the changes in the body mass index of the Brazilian, with the ability to examine heterogeneity by sex and age. The primary aim of the model is to compare intervention scenarios to modelled business as usual (BAU), and can provide outputs such as mortality rates, morbidity rates, life years gained, and disease incidence. Additionally, other outputs such as Quality-Adjusted Life Years (QALYs) and direct and indirect costs of disease may be incorporated to the model.

The conceptual structure of the combination of model is shown below in Figure 1.

**Figure S1. Conceptual structure of the model:**

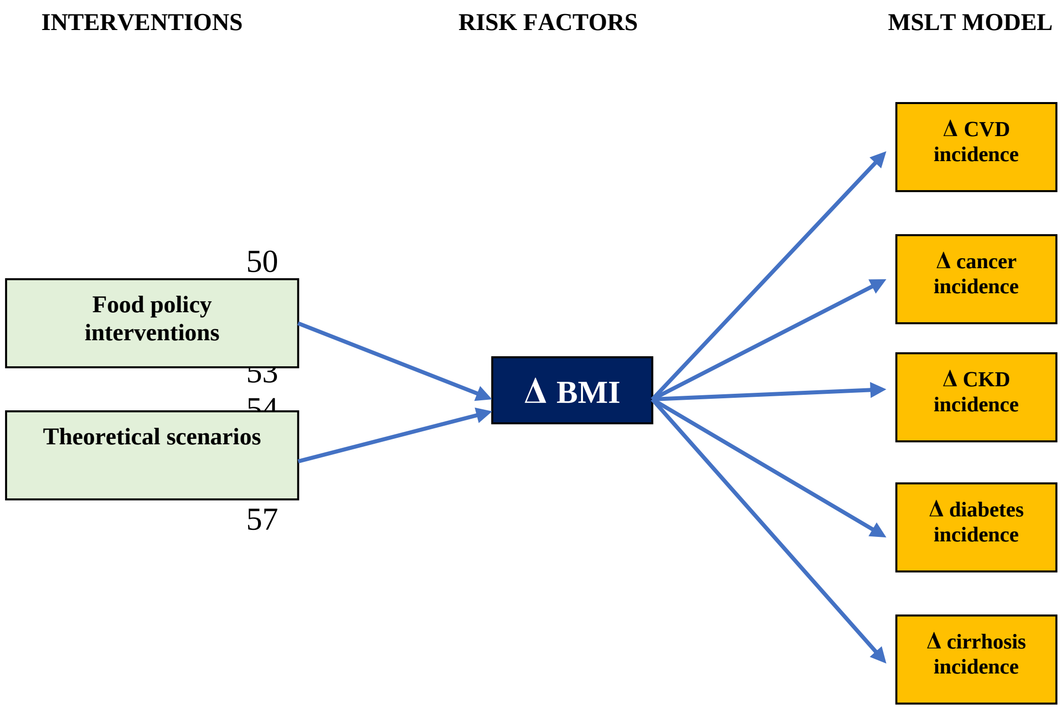


In the future, the existing framework of the model can be completed to also assess dietary interventions through selected foods (fruit and vegetables) and nutrients (sodium and fats), which have epidemiological associations with diseases as reported in the Global Burden of Disease (GBD)(2).

This is a multistate life table (MSLT) macrosimulation model, as PRIME-Time and BODE3 (3)(4). The modelling is carried out for multiple cohorts in parallel by sex sub-populations alive in 2019, for each five-year age group, but can be adapted in the future for different subpopulations. Hence, the model averages or expected values for each cohort. For example, the percentage of each starting sex by age group in each BMI category parametrized using the relative risks of disease outcomes associated with overweight and obesity to generate the population impact fraction (PIF at baseline and in counterfactual scenarios).

**BMI data**

Trends in BMI change were modelled from the Vigitel - Surveillance of risk and protective factors for chronic diseases by telephone survey, using microdata from 2006 to 2019 (5). The business-as-usual (BAU) scenario was estimated as the stability of the rate of increase in BMI through the next 10 years (Tables 1 and 2) and the additional counterfactual scenarios in the analysis consider different changes in which the rate of increase is reduced, if the prevalence of overweight in 2019 remains and if the overweight prevalence among adults was reduced. The average BMI and its standard deviations were estimated for the specific age-groups (20-25, 25-30, 30-35, 35-40, 40-45, 45-50, 55-60, 60-65, 65-70, 70-75, 75-80 and 85+ years) assuming a log-linear distribution in the population.

**Disease modelling - Model structure**

**Life-table analysis**

The model is composed by an overall life table and multiple disease state life tables that are mathematically linked to the main life table. In the baseline or BAU model, the Brazilian population is projected out into the future through all-cause and disease-specific expected trends in incidence, case fatality and mortality. Table 3 includes all the diseases incorporated in this MSLT model and their related specific dietary risk factors, as used in the Global Burden of Disease Study.

The model is a proportional multi-state life-table model, which means that all individuals still alive in each cycle of the model are represented in the main life table, in which age-specific all-cause mortality and morbidity rates are applied in each cycle until the age of 110 years.

In parallel, multiple disease states are modelled independently, in disease-specific life tables. Within these tables, the disease incidence rates, remission and case-fatality rates are modelled. The disease specific life tables have both BAU and intervention models, so that the incidence rates are changed based on population impact fractions (PIFs), allowing the estimation of differences in disease-specific mortality and morbidity rates which are then summed across all parallel disease states (as represented in Figure 2), and added or subtracted to the all-cause mortality and morbidity rates in the main life table.

**Figure S2. MSLT disease states**

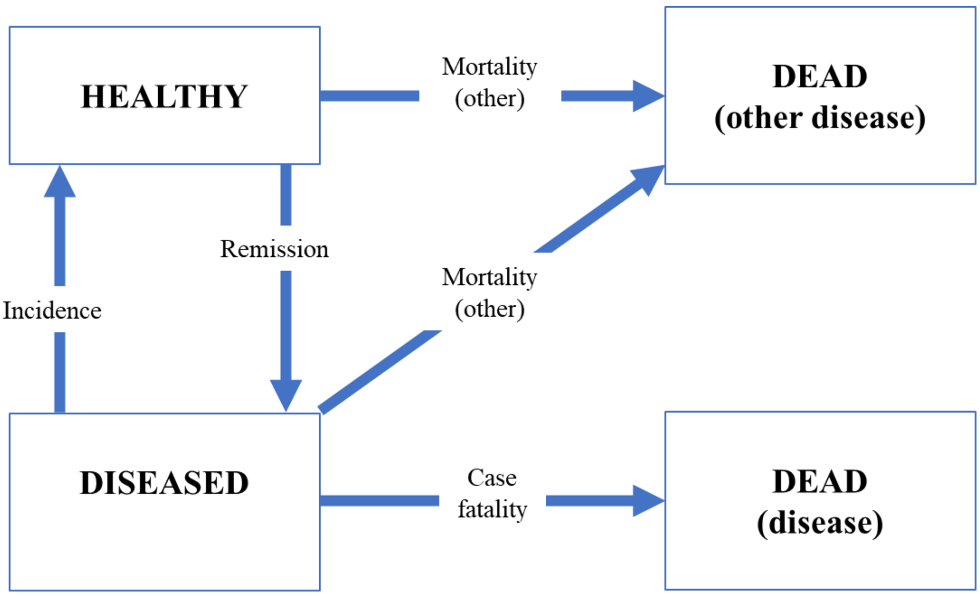


The health impacts of counterfactual scenarios (such as simulated interventions) are achieved by changing risk factors (in this case, BMI) which, in turn, change disease incidence. Therefore, the model shifts risk factor distributions so that the changes are reflected in the resulting PIFs.

As changes in risk factors is normally not associated with immediate or rapid changes in disease incidence, the model also incorporates time lags reflecting the average change in risk factor in a past window of exposure. For example, it was considered that impacts in BMI may affect cardiovascular disease incidence within 5 years, while the time lag for cancers is at least 10 years but might last up to 30 years.

The key assumptions the MSLT are that: the distributions of each risk factor area considered as independent of other risk factors; the incidence rate for each disease is independent of other diseases; and the disease case-fatality and remission rates are independent of those for other diseases.

### Model inputs

The main population parameter included in the model is the population size and the epidemiological parameters such as disease incidence, prevalence, and mortality rates by 5-year age groups for each sex were obtained from the GBD study for Brazil in 2019. The final disease parameters were adjusted using DISMOD II (6), together with the estimation of disease case fatality and remission.

https://doi.org/10.1186/1478-7954-1-4
